# Levels of evidence supporting recommendations in clinical practice guidelines: meta-epidemiological study

**DOI:** 10.1101/2025.08.11.25333132

**Authors:** Kim Boesen, Sarah Louise Klingenberg, Christian Gluud

## Abstract

In this research letter, the levels of evidence supporting recommendations in clinical practice guidelines is assessed. It is a meta-epidemiological study of large-scale guideline assessments.

## Introduction

Clinical practice guidelines are tools to standardize and implement the best available evidence into clinical care. Adherence to clinical guidelines has been associated with improved clinical care.^1^ However, only 11% of recommendations in American cardiology guidelines in 2009 was supported by the highest level of evidence, i.e., multiple randomized clinical trials or systematic reviews, and half the recommendations were supported by the lowest level of evidence such as expert opinion.^2^ Subsequent assessments across other specialties have been conducted, but a global estimate on the proportion of guideline recommendations supported by high level evidence is lacking. It is also unknown whether there is a trend of improvement, stagnation, or worsening over time in the overall evidence base of guideline recommendations.

## Methods

A meta-epidemiological study on the levels of evidence supporting recommendations in clinical guidelines was conducted (protocol: https://doi.org/10.5281/zenodo.11144588). MEDLINE Ovid and Web of Science were searched and records were screened and ranked according to relevance using an open-source machine learning tool, which was trained with 44 relevant and two irrelevant publications. Assessments of >1000 recommendations irrespective of topic and methods were included. The proportion of recommendations supported by the highest level of evidence (according to the method used) and the proportion of the strongest recommendations supported by the highest level of evidence were extracted. A weighted mean proportion, excluding overlapping and ineligible guidelines, was calculated. To assess changes over time, results were categorized as ‘improvement’, ‘no change’, ‘worsening’, or ‘unclear’. Detailed methodology in the Appendix.

## Results

The search returned 149,493 records. We stopped screening early after 360 records due to many relevant hits, at which 115 were included for further assessments. Ninety records were excluded yielding a total of 25 eligible studies (**Figure 1**). The 25 included studies were published between 2007 and 2025, covering 1250 guidelines (range 10 to 168 per study) and 57,166 recommendations (range 1070 to 4289). The weighted mean proportion of recommendations supported by the highest level of evidence was 16% (range 3% to 37% per study) (**Figure 2**). The median proportion (on study level) of recommendations supported by the highest level of evidence (7 studies; 476 guidelines, 21,182 recommendations) was 21% (range 14% to 57%). Changes over time in the levels of evidence were reported in 11 studies (569 guidelines, 23,831 recommendations) and categorized as ‘no change’ (n=8) and ‘worsening’ (n=3), and ‘improvement’ (n=0). See Appendix and Full Dataset for details.

**Figure 1.**
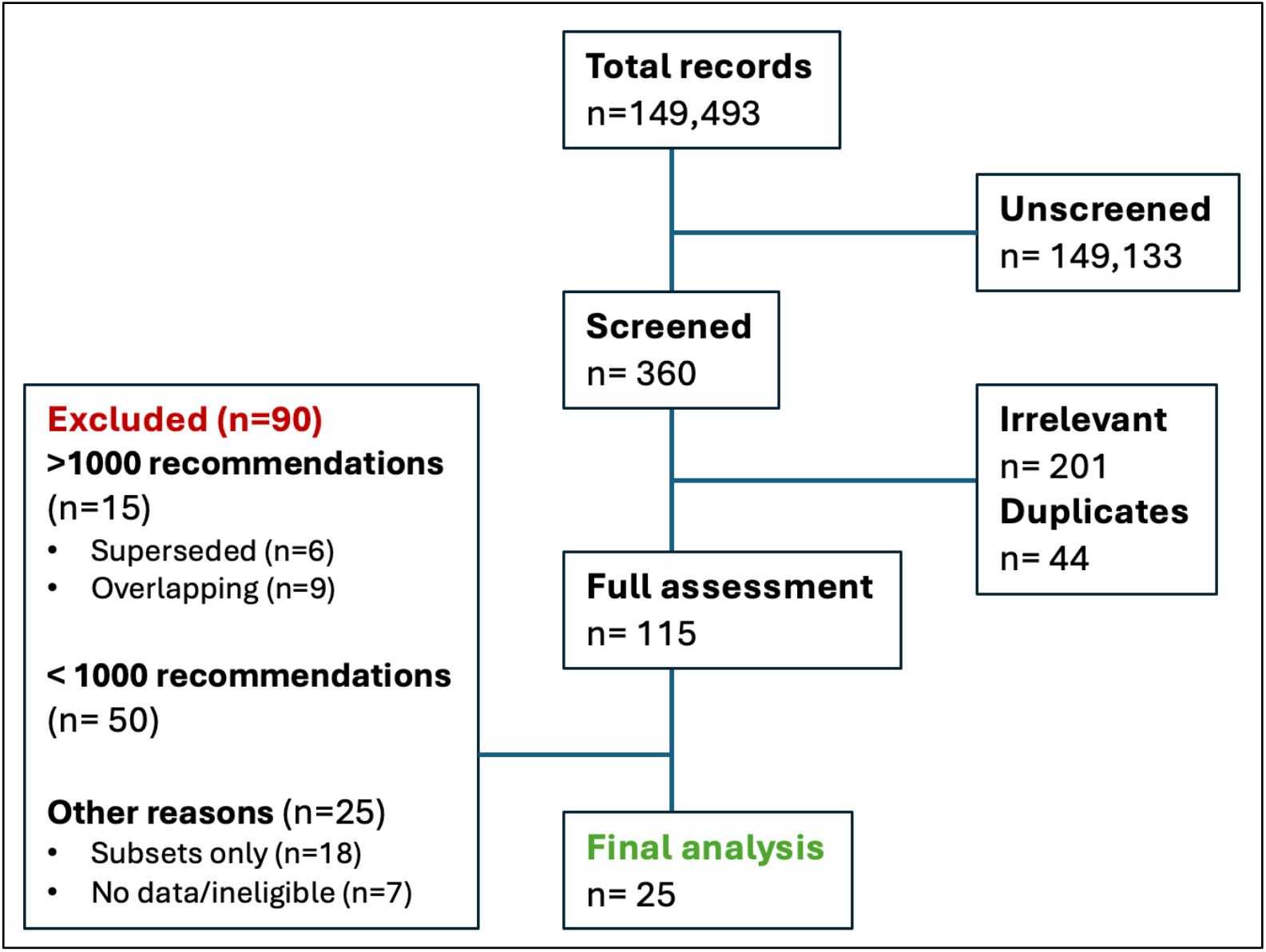
Prisma flowchart

**Figure 2.**
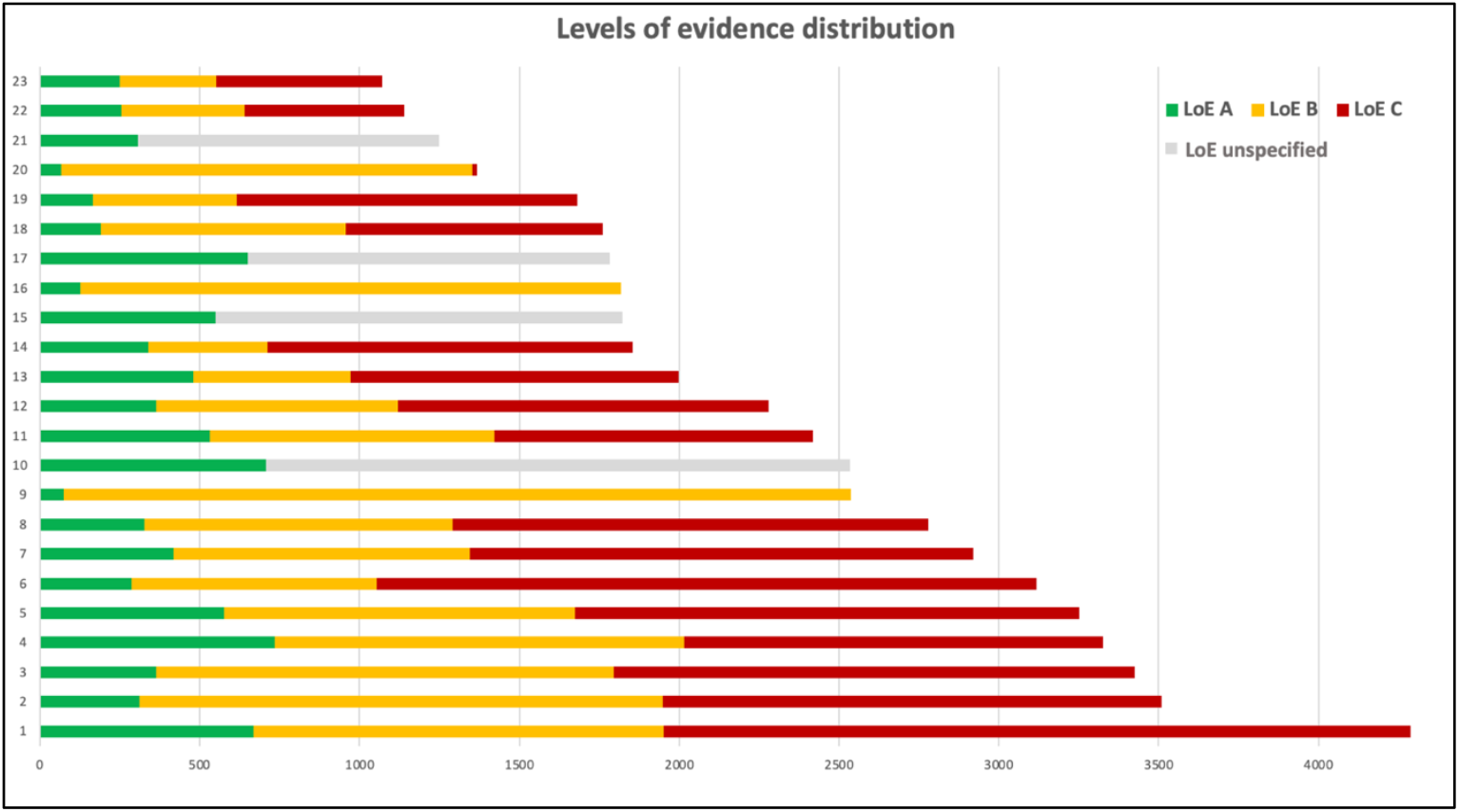
Levels of evidence distribution in included studies 1: Cardiology (Europe, 2023). 2: Cardiology (US, 2019). 3: Interventional medicine (2014). 4: Cardiology (China, 2023). 5: Primary care (2017). 6: Critical care (2018). 7: Infectious diseases (2021). 8: Gastroenterology (US, 2025). 9: Oncology (palliative care, 2012). 10: Gastroenterology (China, 2024). 11: Finnish guidelines (2007). 12: Anesthesiology (2021). 13: Scottish guidelines (2014). 14: Infectious prevention control (2020). 15: Oncology (Europe, 2024). 16: Oncology (Supportive care, 2021). 17: Oncology (systemic treatment, 2019). 18: Endocrinology (2012). 19: Gynecology-obstetrics (UK, 2014). 20: Oncology (hematology, 2021). 21: Gynecology-obstetrics (Canada, 2016). 22: Gastroenterology (liver diseases. 2012). 23: Gastroenterology (inflammatory bowel disease, 2013). Two assessments did not report eligible data. LoE (Levels of evidence), A (usually several randomized clinical trials or systematic reviews), B (usually single randomized clinical trials or observational research), C (usually expert opinion, consensus documents, or best practice). See Dataset and Appendix for details.

## Discussion

This assessment highlights the poverty of evidence supporting clinical guidelines and their recommendations. The level of evidence did not improve substantially in the 11 studies assessing whether the distribution of recommendations changed over time. Several studies noticed that the absolute number of recommendations increased but the distribution in the levels of evidence did not improve.^3,4^ This indicates that newly introduced recommendations are often of low-level evidence, and such recommendations have lower durability and are more frequently removed in updated guidelines.^5^

This study was limited to large-scale assessments using various methods (Appendix). Whether smaller-scale assessments report very different results is unknown. Some classification systems can be misleading since evidence from randomized clinical trials or meta-analyses by default is labelled as highest level, and data from single trials is labelled as lower-level evidence. Solitary, well-conducted trials may be more informative and reliable than meta-analyses of unreliable trials. Some societies have updated their methods to mitigate such limitations.^6^

It is unknown what proportion of highest-level evidence recommendations is desirable and realistic. The global 16% proportion may be useful as an index measure to define specialty-oriented target proportions and to focus on the distribution of evidence instead of ever-increasing number of recommendations.

## Data Availability

The full Dataset (including references to included studies) is available on Zenodo (10.5281/zenodo.16752132)

https://doi.org/10.5281/zenodo.16752132

## Acknowledgements

The Copenhagen Trial Unit for providing salaries for the authors. KB coined the idea, extracted data, and wrote the first draft. SLK conducted the search strategy. All authors revised and agreed on the final manuscript.

