## Appendix for "Levels of evidence supporting recommendations in clinical practice guidelines: meta-epidemiological study"

### Table of content

| | |
|---|---|
| Methods (supplementary) | Page 2 |
| Results (supplementary) | Page 4 |
| References | Page 9 |

### Methods (Supplementary)

### Search strategies

**MEDLINE ALL Ovid (1946 to 18 June 2025) (103684 hits)**

1. *Practice Guidelines as Topic/

2. ((evidence* or "quality of" or "evaluation of" or "review of" or "analysis of") and (guideline* or recommendation* or bulletin* or practice*)).ti.

3. 1 or 2

**Science Citation Index Expanded (1900 to 18 June 2025) and Conference Proceedings Citation Index – Science (1990 to 18 June 2025) (Web of Science) (59448 hits)**

TI=((evidence* or "quality of" or "evaluation of" or "review of" or "analysis of") and (guideline* or recommendation* or bulletin* or practice*))

### Screening

#### ASReview configuration

ASReview's default configuration was used: feature extraction technique (TF-IDF), classifier (Naive Bayes), query strategy (maximum), and balance strategy (dynamic resampling (double)).

#### Training of algorithm

We had knowledge of 44 published relevant guidelines assessments before title/abstract screening.[1-44] These references were used to supervise the ASReview algorithm. Two records were randomly selected as irrelevant records.[45,46]

### Differences between protocol and final report

The study protocol was preprinted 8 May 2024 (https://zenodo.org/records/11144588).

#### Inclusion criteria

We specified the inclusion criteria due to the large number of retrieved relevant studies to limit our assessment to the largest published studies.

- A minimum of 1000 clinical recommendations. This was an arbitrary threshold introduced to limit the number of assessments in our final analysis.
- Overlapping studies were excluded to avoid unit-of-analysis error and double-counting of guidelines and recommendations. We included those studies assessing the largest number of recommendations.
- Superseded assessments, i.e. assessments that had been updated, were excluded from the final analysis to avoid double-counting.
- Studies assessing only select subsets of recommendations from particular guidelines were not included in the final analysis.
- Publications reporting on the development of specific guidance were not included.

**ASReview screening stopping rule**

A stopping rule of 100 consecutive records was prespecified, which was based on previous case studies. We changed the screening strategy due to the large number of relevant records and stopped after 360 records. The recall curve was clearly plateauing (eFigur 1) and we judged we had saturated our sample of eligible assessments at this stage.

**Comparisons over time**

We prespecified to report the changes in proportions of high-level recommendations over time. Due to the differences in how the various studies assessed change (some compared proportions over time, changes in proportions over time, or evidence for individual recommendations) and how this was measured and reported (crude proportion rates, or different test statistics), we opted to classify the individual studies into four categories and descriptively report the results:

- Improvement (a positive change in the evidence base)
- No change (no change in the evidence base supporting recommendations)
- Worsening (lower proportion of high-level recommendations)
- Unclear or not reported

### Results (Supplementary)

#### Screening recall curve

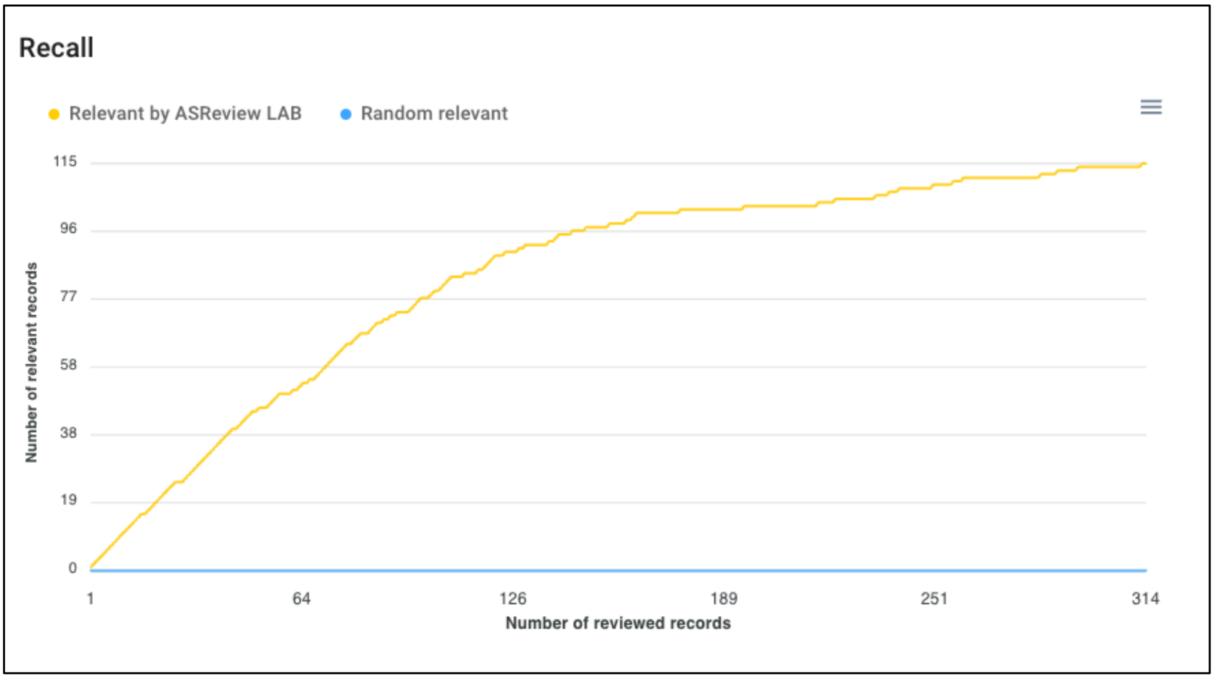


Screenshot from ASReview. The yellow line depicts the recall curve showing the number of screened records (x-axis) compared to the number of included records (y-axis).

#### Overlapping cardiology studies

We included the largest assessments of the American College of Cardiology/American Heart Association (by Bevan et al.[23]) and European Society of Cardiology (by Tantawy et al.[43]). The remaining overlapping assessments were excluded from the analysis.

| **Study** | **ACC/AHA** | **ESC** | **Proportion LoE A** |
|---|---|---|---|
| Tricoci (2009) | 16/1973 | N/A | 11 |
| Faranoff (2019) | 26 / 2930 | 25 / 3399 | 9 / 14 |
| Bevan (2019) | **28 / 3509** | N/A | 9 |

| | | | |
|---|---|---|---|
| DuBrose-Briski (2019) | 16/ 2713 (2008-2012) and 19 / 1998 (2013-2017) | N/A | 13 (2008-2012) and 11 (2013-2017) |
| Van Dijk (2019) | N/A | 27 / 3531 | 13 |
| Caldeira (2020) | N/A | 23 / 3091 | 12 |
| Boriani (2023) | N/A | 50 / 6972[a] | 16 |
| Canepa (2023) | N/A | 18 / 1822[b] | N/A |
| Tantawy (2023) | N/A | **37 / 4289** | 16 |
| Milbradt (2023) | N/A | 29 / 1339[c] | 24 |
| Gomes (2025)d | 29/NA | 47/NA | 8/19 |

ACC (American College of Cardiology), AHA (American Heart Association), ESC (European Society of Cardiology), LoE (Levels of evidence). a) Unit-of-analysis concern and risk of double counting. b) Assessed only six guidelines in three updates. c) Only pharmacological recommendations. d) Number of recommendations/levels of evidence not clearly reported

**Levels of evidence systems**

American Heart Association/American College of Cardiology (**AHA/ACC**)

| | |
|---|---|
| Level of evidence A | Evidence from multiple randomized trials or meta-analyses. |
| Level of evidence B | Evidence from a single randomized trial or non-randomized studies. |
| Level of evidence C | Based on expert opinion, case studies, or standards of care. |

Grading the quality of evidence and the strength of recommendations (**GRADE**)

| | |
|---|---|
| High | We are very confident that the true effect lies close to that of the estimate of the effect. |
| Moderate | We are moderately confident in the effect estimate: the true effect is likely to be close to the estimate of the effect, but there is a possibility that it is substantially different. |
| Low | Our confidence in the effect estimate is limited: the true effect may be substantially different from the estimate of the effect. |

| | |
|---|---|
| Very Low | We have very little confidence in the effect estimate: the true effect is likely to be substantially different from the estimate of effect. |

Strength of Recommendation Taxonomy (**SORT**)

| | |
|---|---|
| A | Recommendation based on consistent* and good quality† patient-oriented evidence |
| B | Recommendation based on inconsistent or limited quality patient-oriented evidence |
| C | Recommendation based on consensus, usual practice, opinion, disease-oriented evidence or case series |

Scottish Intercollegiate Guideline Network (**SIGN)**

| | |
|---|---|
| 1++ | High quality meta-analyses, systematic reviews of RCTs, or RCTs with a very low risk of bias |
| 1+ | Well conducted meta-analyses, systematic reviews, or RCTs with a low risk of bias |
| 1- | Meta-analyses, systematic reviews, or RCTs with a high risk of bias |
| 2++ | High quality systematic reviews of case-control or cohort studies |
| 2+ | High quality case-control or cohort studies with a very low risk of confounding or bias and a high probability that the relationship is causal |
| 2- | Case-control or cohort studies with a high risk of confounding or bias and a significant risk that the relationship is not causal |
| 3 | Non-analytic studies, e.g., case reports and case series |
| 4 | Expert opinion |

National Comprehensive Cancer Network (**NCCN**) method:

| | |
|---|---|
| I | High level of evidence such as randomized controlled trials with uniform consensus |
| IIA | Lower level of evidence with uniform consensus |
| IIB | Lower level of evidence without uniform consensus but no major disagreement |
| III | Any level of evidence but with major disagreement |

Mitchell (2020) method:

| | |
|---|---|
| Level 1 | meta-analyses, systematic reviews of randomized controlled trials (RCTs) |
| Level 2 | well-designed RCTs, strong recommendation from high-quality evidence |
| Level 3 | well-designed non-randomized studies including observational studies (case–control, cohort and cross-sectional), moderate evidence; |
| Level 4 | descriptive studies, expert opinion, low-quality evidence; |
| Level 5 | poor/insufficient evidence, very-low-quality evidence; |
| Level 6 | recommended best practice, good practice point; |
| Level 7 | unresolved issue, no recommendation; |
| Level 8 | legislated requirement. |

Skelin (2020) method:

| Level of evidence | |
|---|---|
| I | Evidence from at least one large randomised, controlled trial of good methodological quality (low potential for bias) or meta-analyses of well-conducted randomised trials without heterogeneity |
| II | Small randomised trials or large randomised trials with suspicion of bias (lower methodological quality) or meta-analyses of such trials or of trials with demonstrated heterogeneity |
| III | Prospective cohort studies |
| IV | Retrospective cohort studies or case–control studies |
| V | Studies without control group, case reports, expert opinions |

Prusova (2014) method:

| | Classification scheme for recommendations by the Royal College of Obstetricians and Gynaecologists |
|---|---|
| Pre-December 2007 | |
| A | At least one randomised controlled trial as part of a body of literature of overall good quality and consistency addressing the specific recommendation. |
| B | Requires the availability of well controlled clinical studies but no randomised clinical trials on the topic of recommendations. |
| C | Requires evidence obtained from expert committee reports or opinions and/or clinical experiences or respected authorities. Indicates an absence of directly applicable clinical studies of good quality. |
| D | Recommended best practice based on the clinical experience of the guideline development group. |
| Post-December 2007 | |
| A | At least one meta-analysis, systematic review or randomised controlled trial rated as 1++ directly applicable to the target population and demonstrating overall consistency of results.<br>A systematic review of randomised controlled trials or a body of evidence consisting principally of studies rated as 1 + directly applicable to the target population and demonstrating overall consistency of results. |
| B | A body of evidence including studies rated as 2++ directly applicable to the target population, and demonstrating overall consistency of results.<br>Extrapolated evidence from studies rated as 1++ or 1+. |
| C | A body of evidence including studies rated as 2 + directly applicable to the target population and demonstrating overall consistency of results.<br>Extrapolated evidence from studies rated as 2++. |
| D | Evidence level 3 or 4.<br>Extrapolated evidence from studies rated as 2+. |
| E | Recommended best practice based on the clinical experience of the guideline development group. |

Ghui (2014) method:

| | |
|---|---|
| I | Evidence obtained from at least one properly randomised controlled trial |
| II-1 | Evidence from well-designed controlled trials without randomisation |
| II-2 | Evidence from well-designed cohort (prospective or retrospective) or case–control studies, preferably from more than one centre or research group |
| II-3 | Evidence obtained from comparisons between times or places with or without the intervention. Dramatic results in uncontrolled experiments (such as the results of treatment with penicillin in the 1940s) could also be included in the category |
| III | Opinions of respected authorities, based on clinical experience, descriptive studies, or reports of expert committees |
